# Young women’s knowledge and attitudes towards cervical cancer screening at a selected higher education institution in Lesotho

**DOI:** 10.1101/2021.10.11.21264801

**Authors:** Isabel Nyangu, Tanki Moteane

## Abstract

**Background:** In Lesotho, cervical cancer is the most common female cancer and leading cause of death amongst women. In 2019, the annual number of new cancer cases was 477 and 346 women died from cancer related complications in Lesotho.

**Aim:** The aim of this study was to assess the knowledge and attitudes of young women towards cervical cancer screening at a selected higher education institution in Lesotho.

**Methods:** A quantitative cross-sectional research design was used to collect data using an interviewer administered semi-structured questionnaire from 80 young women who were randomly selected. Permission to conduct the study was sought and granted from relevant authorities. Informed consent was sought from the respondents who were identified using codes and participated voluntarily.

**Results:** Fifty percent (n=40) of the women did not know cervical cancer screening tests, 70% (n=56) did not know about the frequency of screening, whilst 75% (n=60) knew that human papilloma virus (HPV) vaccine is be used to prevent cervical cancer. Additionally, 95% (n=76) had not screened for cervical cancer, 65% (n=52) perceived cervical cancer screening as painful, and 95% (n=76) needed more information on cervical cancer screening.

**Conclusions:** The practice of cervical cancer screening is low and there is a high need for health education and incorporating regular cervical cancer screening in health care services to increase the uptake amongst young women. Many participants were hesitant to screen as they perceived the procedure to be painful and this suggests the need for reassurance and counselling on cervical cancer screening techniques.

## Introduction

Globally, cervical cancer is the fourth common type of cancer accounting for 6.6 % of all female cancers,[1] with China and India together contributing more than a third of the global cervical cancer burden (106 000 cases and 48 000 deaths in China and 97 000 cases and 60 000 deaths in India),[2]. Cervical cancer is one of the most preventable and curable forms of cancers through early detection and treatment. Unfortunately, in many parts of Africa, cervical cancer was previously not identified or treated until it had reached an advanced stage due to insufficient access to health care services, effective screening and early treatment,[3].

Cervical cancer has been steadily increasing in sub-Saharan Africa, with more than 75 000 cases and 50 000 deaths yearly, and is further exacerbated by HIV,[4]. In southern Africa, 63.8% of women with cervical cancer were living with HIV, as were 27·4% of women in eastern Africa,[5]. The highest incidence was reported in Eswatini, with approximately 6.5% of women developing cervical cancer before the age of 75 years,[2]. The incidence rate of cervical cancer in South Africa was reported between 22.8 and 27 per 100 000 women as compared with the global average of 15.8, and annually there were approximately 6 000 new cases reported with 3 000 deaths,[6].

With the overall burden of cervical cancer on most African countries expected to surge over the next decades, Lesotho has taken few initiatives to reduce its impact, by introducing free cervical screening program using visual inspection of the cervix with acetic acid (VIA) and Pap test,[7]. In Lesotho, cervical cancer was the leading cause of death among women with an incidence rate of 27.8/100 000 in 2012,[7] and this was attributed to the high HIV incidence and prevalence. Current estimates indicated that every year in Lesotho, 477 women were diagnosed with cervical cancer and 346 died from the disease, particularly in women aged between 40 and 55 years,[7]. Additionally, women present themselves at health centres only when they are sick and this makes it difficult for the health personnel to diagnose the infection at its early and curable stages,[8].

The incidence and prevalence rate of cervical cancer among women in Lesotho continues to increase, with an average of 18 confirmed cases of cervical cancer per month. Despite the availability of free cervical cancer screening services provided in Lesotho, uptake remains limited. This study therefore sought to assess young women’s knowledge and attitudes towards cervical cancer screening in Roma, Lesotho

## Methodology

A quantitative cross-sectional survey was conducted with a sample of 80 young women who were randomly sampled. Data was collected using an interviewer administered semi-structured questionnaire in the months of June-July 2021. A pilot study using 15 respondents who were not included in the study was conducted to ensure validity and reliability of the data collection tool. The questionnaire had three sections which collected information on the respondents’ demographics, knowledge, and attitudes towards cervical cancer screening. Permission to conduct this study was sought and granted from relevant authorities. Codes were used to identify the participants and data was collected in a private and quiet environment. Respondents provided written consent and voluntarily took part in this study. They were allowed to withdraw from the study without any repercussions. They were allowed to ask clarity seeking questions before responding to the questions. Upon completion of the study, the final report was availed to the relevant authorities and library at the selected higher education institution.

## Results

Thirty percent (n=24) of the respondents were aged 18-20 years, 45% (n=36) were aged 21-25 years, 20% (n=16) were aged 26-30, and 5% (n=4) were aged 31-35 years. Sixty (75%) were single, 25% (n=20) were married, and all of them were sexually active and Christians. Forty-eight (60%) were from the rural areas, while 40% (n=32) were from the urban areas.

### Knowledge on Cervical cancer screening

Most of the respondents (75%; n=60) knew that Human Papilloma Virus (HPV) causes cervical cancer. Some of the respondents (40%; n=32), knew that the signs and symptoms of cervical cancer included coital pain, 30% (n=24) knew about post-coital bleeding, 20% (n=16) knew about post-menopausal bleeding, and only 10% (n=8) knew that cervical cancer can be asymptomatic. Half (50%; n=40) of the respondents knew about the screening tests for cervical cancer. Some of them (45%; n=36) had information that Pap smear could be used to diagnose cervical cancer, and 5% (n=4) knew that VIA could be used for screening.

Majority (70%; n=56) of the respondents did not know how often one should screen for cervical cancer, 20% (n=16) said that an individual should screen every year, and 10% (n=8) pointed that screening should be done every 3 years. When asked who is eligible for cervical cancer screening, majority (55%; n=44) of the respondents did not know, 30% (n=24) pointed women whose age was greater than 21 years, and 15% (n=12) pointed that all sexually active women should screen.

Most (75%; n=60) of the respondents suggested that HPV vaccine was the preventive measure against cervical cancer, and 15% (n=12) pointed that regular screenings are the preventive measure. Figure 1 below illustrates the knowledge in more detail.

**Figure 1:**
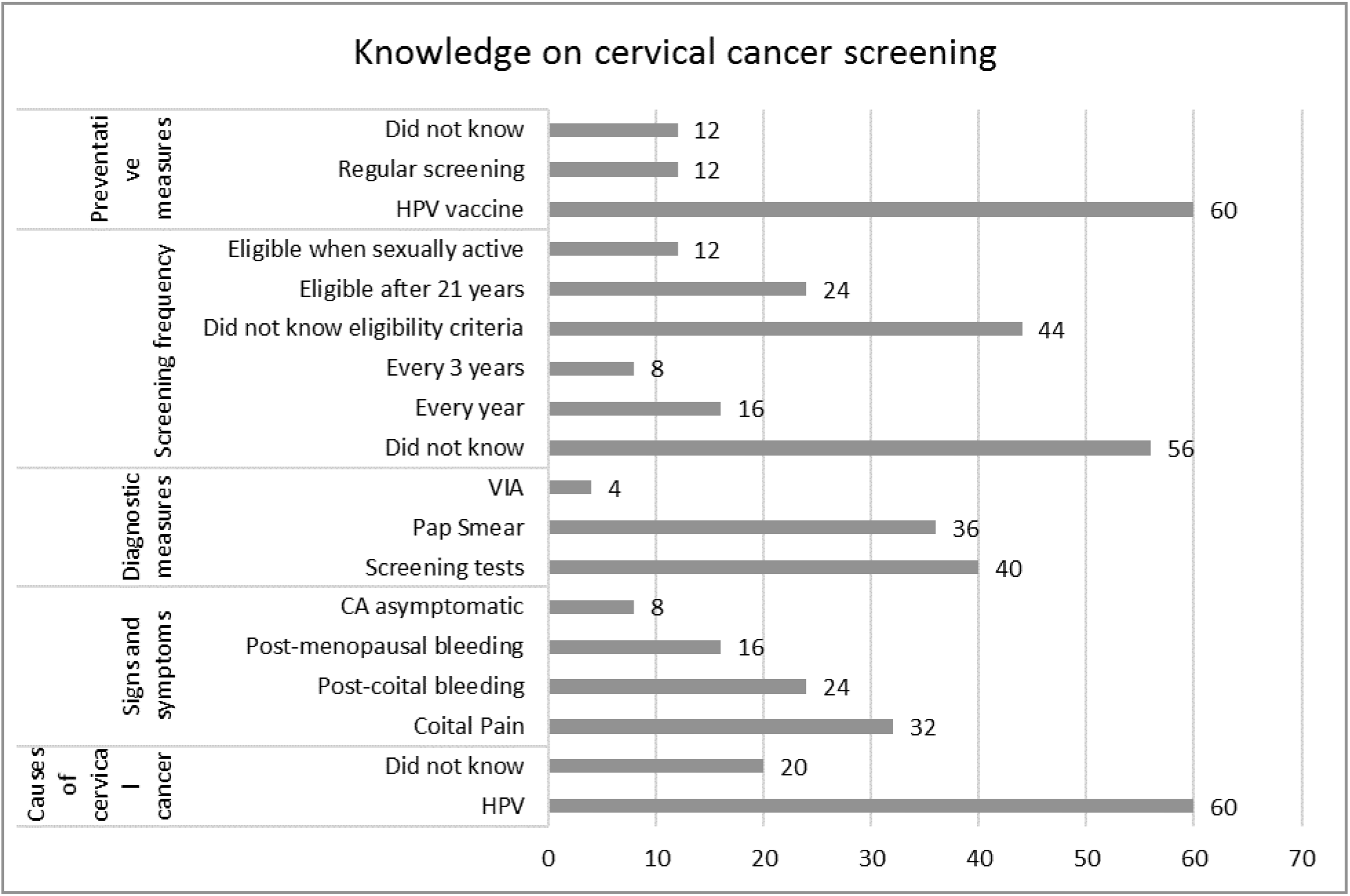
Knowledge on cervical cancer screening.

### Attitudes towards cervical cancer screening

Most (95%: n=76) of the respondents had not yet been screened for cervical cancer, while 5% (n=4) had screened. Majority (65%: n=52) of the respondents thought that the technique used for cervical cancer screening was painful, while 35% (n=28) said it caused discomfort. Majority (80%: n=64) of the respondents had not screened for cervical cancer due to lack of healthcare facilities in their homes, 15% (n=12) reported that they had never been asked by a doctor, while 5% (n=4) reported to have felt no need to screen. Majority (75%: n=60) of the respondents preferred to be screened by doctors whilst 25% (n=20) preferred to be screened by nurses. Majority (95%: n=76) of the respondents needed more information on cervical cancer screening.

## Discussion

The age group of 21 to 25 years (45%) was the largest which is contrary to the age group 26 to 30 years (50%) in a study conducted in Niger,[9]. All the respondents were Christians. Engaging religious groups in educating and encouraging their members about cervical cancer screening can also help in reaching women,[10].

Respondents (75%) knew that HPV was a risk factor of cervical cancer. This is in in line with a study by McBride and Singh,[11] in which more than 70% of the respondents were aware that HPV could cause cervical cancer but in contrast to a study by Vishwakarma et al.,[12] where only 32% of subjects knew about HPV. A few of the respondents were aware that cervical cancer can be asymptomatic, present as irregular menstrual bleeding, and present as vaginal bleeding after intercourse. This is less as compared to the study by Vishwakarma et al.,[12] and Urasa and Darj,[13] where more respondents were aware.

Pap smear was the most common screening test known among respondents. This is very similar to a study carried by Owoeye and Ibrahim,[9] in which 44.9% of the respondents knew about a pap smear test. However, this was very low compared to 88.4%, 83%, and 82% of the respondents that knew in studies by Shah et al.,[14], Mutyaba et al.,[15] and Vishwakarma et al.,[12], respectively. Other screening methods such as cytology-based and HPV DNA which are commonly used in developed countries are not available in Lesotho due to financial and technical constraints.

Cervical cancer screening through Pap smear could be very effective in detecting cervical neoplasia at a primordial stage, and this could be precipitated by a very effective treatment. Only 30% of the respondents knew that screening should begin at 21 years, which is comparable to 46% and 54.1% of respondents that knew in studies by Vishwakarma et al.,[12] and Shah et al.,[14], respectively. Majority of the respondents in this study knew that HPV vaccination was a preventive measure against cervical cancer, and this is a similar finding to a study that reported that 82.7% of the participants pointed HPV as a preventive measure,[16]. Similarly, majority of the respondents preferred doctors over nurses in performing the Pap smear tests. This is similar to a study by Shekhar et al.,[17] in which 71% the respondents preferred doctors over nurses, because they perceived cervical cancer screening to be a doctors’ procedure.

The higher mortality of cervical cancer in Lesotho can be attributed to late presentation of the cancer at health facilities. In this study, only 5% of the respondents had a Pap smear. This is very similar to 5.7% uptake in Nnewi,[18] and less as compared to 8.3% among Nigerian women,[19]. Another study conducted in Niger reported an 11.6% uptake,[9]. In a study carried out in Nigeria, the uptake was 8.7%,[20], and 8.5% in a study conducted in Ghana,[21]. In contrast, the uptake was 84% amongst Chinese American women in the United States,[22], whilst more than 80% of the adult females in United States had a Pap smear during the preceding 3 years,[23]. In majority of developing countries, particularly Lesotho, implementing such measures of coordinated intervention has seemed far impractical.

Poor screening uptake can be attributed to limited healthcare facilities in rural areas which are mostly found in highland regions of the country. In this study, majority of the respondents came from rural areas. This has also been reported in other studies where uneven distribution of medical facilities in the country was the reason for poor uptake,[24], and long distances to such facilities also affected uptake,[16]. In contrast, other reasons for poor cervical cancer screening was due lack of knowledge about the availability of screening, and culturally-influenced reluctance to undergo cervical smear tests,[25].

Respondents who had Pap smears experienced pain (65%) and discomfort (35%), which are deemed as misconceptions by other researchers,[26,27]. The respondents were however keen to obtain more information about cervical cancer screening and this is similar to a study by Imam et al.,[28] where 95% showed interest in obtaining more information.

### Recommendations

Health education on cervical cancer screening should be given extensively by the all healthcare professionals to strengthen women’s knowledge and encourage them to participate in the uptake of cervical cancer screening. Extensive health education on cervical cancer screening needs to be enhanced through media outlets such as radios. Interventions should focus on detailing the nature of the sample and teaching women relaxation techniques so that they can cope with pain and discomfort. Healthcare professionals should only reinforce positive behaviours, so that more women can screen for cervical cancer without fear or any attitude.

## Strengths and limitations

The study was limited to the knowledge and attitudes of young women on cervical cancer screening.

## Conclusion

The study shows that the practice of cervical cancer screening is low and there is a high need for information pertaining to cervical cancer screening. There is a need for incorporating regular cervical cancer screening in health care services to increase the uptake amongst women.

## Data Availability

All data produced in the present work are contained in the manuscript

## Acknowledgements

Special gratitude goes to the National Manpower Development Secretariat for funding this study.

